# OpenSAFELY: impact of national guidance on switching from warfarin to direct oral anticoagulants (DOACs) in early phase of COVID-19 pandemic in England

**DOI:** 10.1101/2020.12.03.20243535

**Authors:** The OpenSAFELY Collaborative, Helen J Curtis, Brian MacKenna, Alex J Walker, Richard Croker, Amir Mehrkar, Caroline E Morton, Seb Bacon, George Hickman, Peter Inglesby, Chris Bates, David Evans, Tom Ward, Jonathan Cockburn, Simon Davy, Krishnan Bhaskaran, Anna Schultze, Christopher T Rentsch, Elizabeth Williamson, William Hulme, Helen I McDonald, Laurie Tomlinson, Rohini Mathur, Henry Drysdale, Rosalind M Eggo, Kevin Wing, Angel YS Wong, Harriet Forbes, John Parry, Frank Hester, Sam Harper, Stephen JW Evans, Ian J Douglas, Liam Smeeth, Ben Goldacre

## Abstract

**Background:** Early in the COVID-19 pandemic the NHS recommended that appropriate patients anticoagulated with warfarin should be switched to direct acting oral anticoagulants (DOACs), requiring less frequent blood testing. Subsequently, a national safety alert was issued regarding patients being inappropriately co-prescribed two anticoagulants following a medication change, and associated monitoring.

**Objective:** To describe which people were switched from warfarin to DOACs; identify potentially unsafe co-prescribing of anticoagulants; and assess whether abnormal clotting results have become more frequent during the pandemic.

**Methods:** Working on behalf of NHS England we conducted a population cohort based study using routine clinical data from >17 million adults in England.

**Results:** 20,000 of 164,000 warfarin patients (12.2%) switched to DOACs between March and May 2020, most commonly to edoxaban and apixaban. Factors associated with switching included: older age, recent renal function test, higher number of recent INR tests recorded, atrial fibrillation diagnosis and care home residency. There was a sharp rise in co-prescribing of warfarin and DOACs from typically 50-100 per month to 246 in April 2020, 0.06% of all people receiving a DOAC or warfarin. INR testing fell by 14% to 506.8 patients tested per 1000 warfarin patients each month. We observed a very small increase in elevated INRs (n=470) during April compared with January (n=420).

**Conclusions:** Increased switching of anticoagulants from warfarin to DOACs was observed at the outset of the COVID-19 pandemic in England following national guidance. There was a small but substantial number of people co-prescribed warfarin and DOACs during this period. Despite a national safety alert on the issue, a widespread rise in elevated INR test results was not found. Primary care has responded rapidly to changes in patient care during the COVID-19 pandemic.

## Background

Following the onset of the COVID-19 pandemic and the implementation of “lockdown”, the NHS responded to deliver healthcare services in a manner that minimised risk of transmission of severe acute respiratory syndrome coronavirus 2 (SARS-CoV-2). Most Patients taking the anticoagulant warfarin require frequent blood tests, the International Normalised Ratio (INR) test, potentially increasing their chance of exposure to SARS-CoV-2. NHS England issued guidance in late March 2020 [1] to support local NHS organisations to manage their anticoagulant services; this included identifying people suitable for switching from warfarin to direct oral anticoagulants (DOACs), which require less frequent blood testing. Subsequently, in October 2020, the Medicines and Healthcare products Regulatory Authority (MHRA) issued a safety alert, warning about an increase in the number of people with substantially elevated INR levels observed during the pandemic and also warned that of some people for whom warfarin was inadvertently continued after switching to DOACs [2].

Anticoagulants are prescribed to people at risk of, or for treatment of thromboembolism, which in some cases can result in stroke. Warfarin, a vitamin K antagonist, has been the mainstay of oral anticoagulant treatment for decades. Unlike most medicines, a patient’s specific dose of warfarin is adjusted based upon frequent blood tests which determine their INR, a measurement of blood clotting time. Factors such as changes in diet, alcohol intake, acute illness and concomitant medications can affect blood levels of warfarin and the INR, requiring a temporary increase in frequency of testing. The quality of the anticoagulation control is assessed by the proportion of time in therapeutic range (TTR). DOACs’ (rivaroxaban, dabigatran etexilate, apixaban, and edoxaban) mechanism of action does not alter the INR and therefore people taking DOACs only require less frequent drug safety monitoring (for example renal function), in line with many other commonly-prescribed medications. Nationally, the prescribing of DOACs has increased steadily since their recommendation by NICE for atrial fibrillation in 2012 [3], but they remain more expensive than warfarin.

In late May 2020 NHS England wrote to Clinical Commissioning Groups (CCGs), the local NHS bodies responsible for medicines use, advising that apixaban or rivaroxaban should be prescribed in place of warfarin for people able to change, [4] following a procurement exercise that secured additional stock at reduced prices. Subsequently, in October 2020, the Medicines and Healthcare products Regulatory Authority (MHRA) issued their safety alert about co-prescribing of warfarin and DOACs and elevated INRs [2]; however this document gave no indication of the scale of these problems.

Using a retrospective cohort study design, we set out to: evaluate the proportion and characteristics of prior warfarin users who switched to DOACs, and how many subsequently reverted; identify potentially unsafe co-prescribing of warfarin and DOACs; measure the frequency of INR testing during the pandemic for people taking warfarin, any changes to TTR, and the extent to which elevated INRs were observed. This was conducted as a “proof of concept” for the use of the new OpenSAFELY analytics platform to rapidly understand service impacts during the COVID-19 pandemic and inform support for primary care.

## Methods

### Study design

Prescribing and testing practice was analysed by conducting a retrospective cohort study using data from English NHS general practices.

### Data sources

Primary care records managed by the GP software provider The Phoenix Partnership (TPP) were assessed using OpenSAFELY, a data analytics platform created by our team on behalf of NHS England to address urgent COVID-19 research questions (https://opensafely.org). Open SAFELY provides a secure software interface allowing the analysis of pseudonymized primary care patient records from England in near real-time within the EHR vendor’s highly secure data centre, avoiding the need for large volumes of potentially disclosive pseudonymized patient data to be transferred off-site. This, in addition to other technical and organisational controls, minimizes any risk of re-identification. Similarly pseudonymized datasets from other data providers are securely provided to the EHR vendor and linked to the primary care data. The dataset analysed within OpenSAFELY is based on 24 million people currently registered with GP surgeries using TPP SystmOne software (40% of England’s population). It includes pseudonymized data such as coded diagnoses, medications and physiological parameters, but no free text data. Further details on our information governance can be found on page 17, under information governance and ethics.

### Data processing

We extracted data for all people who were issued with a prescription or had an active repeat prescription for warfarin or DOACs between January 2019 and August 2020, and the relevant dates of each prescription and INR test-related activity. We used warfarin, DOAC and INR codelists from OpenSAFELY [5,6] to identify these activities, with “high INRs” classified as INR results values >8 / ≥8 (as indicated) in line with thresholds relied upon by the MHRA [7]. We identified recorded INR time in therapeutic range (TTR) using the following CTV3 codes ‘YavzQ’, ‘42QE2’ (redundant), ‘Xaa68’.

### People switching from warfarin to DOACs following NHS England guidance

We identified people issued a prescription for warfarin (but no DOACs) between December 2019-February 2020, and assessed how many received at least one DOAC (“switched”) in March-May 2020, or only received warfarin (Figure 1). Of those switching, we counted how many later received warfarin again during this period (“switched back”). Of those remaining on warfarin we counted how many received an INR test, a high INR value, or had a TTR recorded. This analysis was repeated for the subsequent 3-month period (warfarin March-May 2020, switching June-August 2020), and for the corresponding periods in the previous year. Descriptive tables were generated to describe the cohort.

**Figure 1.**
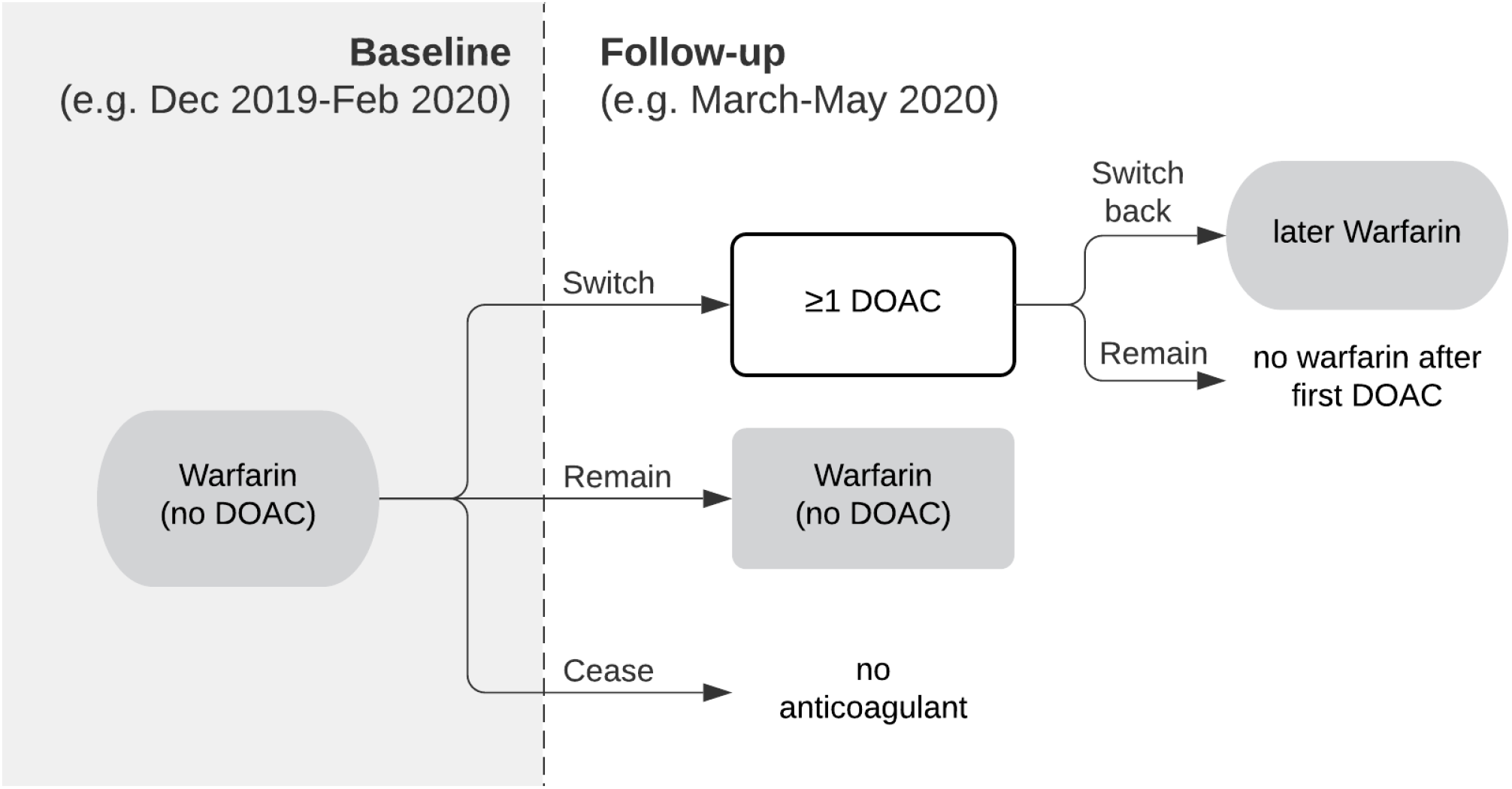
Flow chart illustrating how patients switching from warfarin to DOACs (and those switching back within the same period) were identified based upon prescriptions issued.

We also assessed the overall rate of people starting new DOAC repeat prescriptions each month from January 2019-August 2020 (with “new” defined as people having no DOAC repeat prescription ending within the previous three months). We assessed how many of whom had switched from warfarin, defined as people having a warfarin repeat prescription with an end date in the previous three months. Figures were plotted on a time trend chart.

### Potentially unsafe co-prescribing of warfarin and a DOAC

For each month, January 2019-August 2020, patients were identified who: a) were issued a prescription for DOAC and warfarin on the same day, or b) had a repeat medication initiated for both DOAC and warfarin on the same day. We identified whether these people had at least one of these prescriptions ended the same day. Figures were plotted on a time trend chart.

### INR blood tests during the COVID-19 period

Each month, INR tests and TTR values were identified for people on warfarin (defined as people with warfarin issued within the previous 3 months, and no DOAC issued after the latest warfarin). We plotted the rate per 1000 warfarin patients (and for TTRs, also the rate per 1000 INR tests and the rate per 1000 people tested) on time trend charts.

### Factors associated with switching from warfarin to a DOAC during the pandemic

We investigated the factors associated with switching from warfarin to DOAC using mixed effects logistic regression. We included the variables in Table 1 as factors in the model. We also planned to include the patient’s practice but this was later removed to allow the model to converge. The population was: people aged 18-110 at the start of the follow-up period; registered with a single practice for the six months prior to the follow-up period (to ensure completeness of covariate data); prescribed warfarin at least once in the baseline period; first prescribed warfarin at least 6 months ago (to exclude very recent initiations); not prescribed a DOAC during the baseline period; and prescribed at least one DOAC or warfarin during the follow-up period (to exclude people who ceased treatment or died before receiving any prescription in this period). The outcome was the prescription of at least one DOAC during the follow-up period. Baseline period: 3 months prior to the national “stay at home” order (16 December 2019-15 March 2020); follow-up period: the following 3 months (16 March 2020-15 June 2020).

**Table 1.** Variables used to assess factors associated with switching (present on the first day of the follow-up period unless otherwise stated).

| Factor | Detailed information on coding | Codelist (where applicable) |
| --- | --- | --- |
| Age Group | <65, 65-74, 75+ |  |
| Ethnicity | 6 categories.<br>Ethnicity will be missing for ~30% of people so will not be included in the primary model. | https://codelists.opensafely.org/code-list/opensafely/ethnicity/2020-04-27/ |
| Indices of deprivation (IMD) | Deciles based on residential postcode | Supplied by TPP |
| Sustainability and transformation partnership (STP) membership | STP of current registered practice - modelled as a random effect | STP are NHS administrative regions made up of one or more CCGs<br>Linkage to national STPs carried out by TPP |
| Care / nursing home residence status | Yes/no | Classification supplied by TPP based on patient's residential address at 1 Feb 2020 linked to CQC care home register. |
| Diagnosis of atrial fibrillation | Yes/no; any time prior to follow-up period | https://codelists.opensafely.org/code-list/opensafely/atrial-fibrillation/2020-07-09/ |
| Renal Function | Latest eGFR function (<30, 30-59, ≥60) in the 12 months to the end of the baseline period (or “no evidence” for those with no creatinine recorded in this period). | eGFR calculated from creatinine records.<br>Creatinine clearance is recommended renal function assessment for tailoring DOAC doses for individual people. Automated calculation of CrCl is not yet available in OpenSAFELY and eGFR is used as pragmatic alternative. |
| Recent Renal Function Test | Yes/no; within last 4 months and up to the end of the study period (this conservatively allows tests slightly outside of the recommended 3 month period, and any time during the switching period). | Creatinine test, or:<br>• “451...” Renal function test<br>• “XacUK” eGFR using creatinine (CKD-EPI) per 1.73 square metres |
| Number of INR tests | 0, 1-3, 4-6, or 7+ tests during the baseline period | https://codelists.opensafely.org/code-list/opensafely/international-normalised-ratio-inr/2020-10-22 |
| Length of time on warfarin* | Categorised as:<br>• ≤ 2 years (since 16 March 2018)<br>• ≤ 6 years (since June 2014, date of latest NICE AF guidance [8])<br>• ≤ 8 years (since March 2012, date of first NICE Technology Appraisal for a DOAC for AF [9]) | https://codelists.opensafely.org/code-list/opensafely/warfarin/2020-10-05/ |
|  | • > 8 years |  |
| Previously prescribed DOACs* | Yes/no; any time prior to the baseline period | https://codelists.opensafely.org/codelist/opensafely/direct-acting-oral-anticoagulants-doac/2020-10-05/ |
| Explicitly Recorded contraindication to DOAC | Yes/no; any time prior to the follow-up period | https://codelists.opensafely.org/codelist/opensafely/explicit-contraindication-to-doacs-direct-acting-anticoagulants/2020-10-19/ |

*\*The earliest actual date of an event may be missing due to incomplete patient records (e.g. lost in transfer between practices), but the earliest recorded date can be used as an approximation.*

### Software and reproducibility

Data management was performed using Python 3.8 and SQL, and regression analysis using Stata 16.1. All code for the OpenSAFELY platform, and for data management and analyses for this study, are available for inspection and reuse under open licenses on GitHub (https://github.com/opensafely/anticoagulant-switching-research). All codelists are available for inspection and re-use from https://codelists.opensafely.org/.

### Patient and Public Involvement

Patients were not formally involved in developing this specific study design that was developed rapidly in the context of a global health emergency. We have developed a publicly available website https://opensafely.org/ through which we invite any patient or member of the public to contact us regarding this study or the broader OpenSAFELY project.

## Results

### Patients switching from warfarin to DOACs following NHS England advice

164,000 people were prescribed warfarin between December 2019-February 2020, of whom 12.2% (20,000) were prescribed a DOAC between March-May 2020 (Table 2). This is substantially higher than the previous year (3.5%), and the following three months (4.4%) (Table 2). Of those who switched to a DOAC, 5.8% (n=1,200) also received a subsequent prescription for warfarin (switched back), compared to 4.1% the previous year (Table 2). Of those remaining on warfarin, 80.1% had at least one INR test recorded during March to May compared to 83.7% the previous year, 38.6% had at least one recorded INR TTR (37.9% the previous year), and 0.5% had a high INR (≥8) result (0.4% the previous year; Table 2). During March-May 2020, edoxaban and apixaban were the most commonly selected DOACs (38.1% and 33.5% respectively; Table 3). During June-August 2020, apixaban had increased to 40.3%, with edoxaban dropping to 33.4% (Table 3).

**Table 2.**
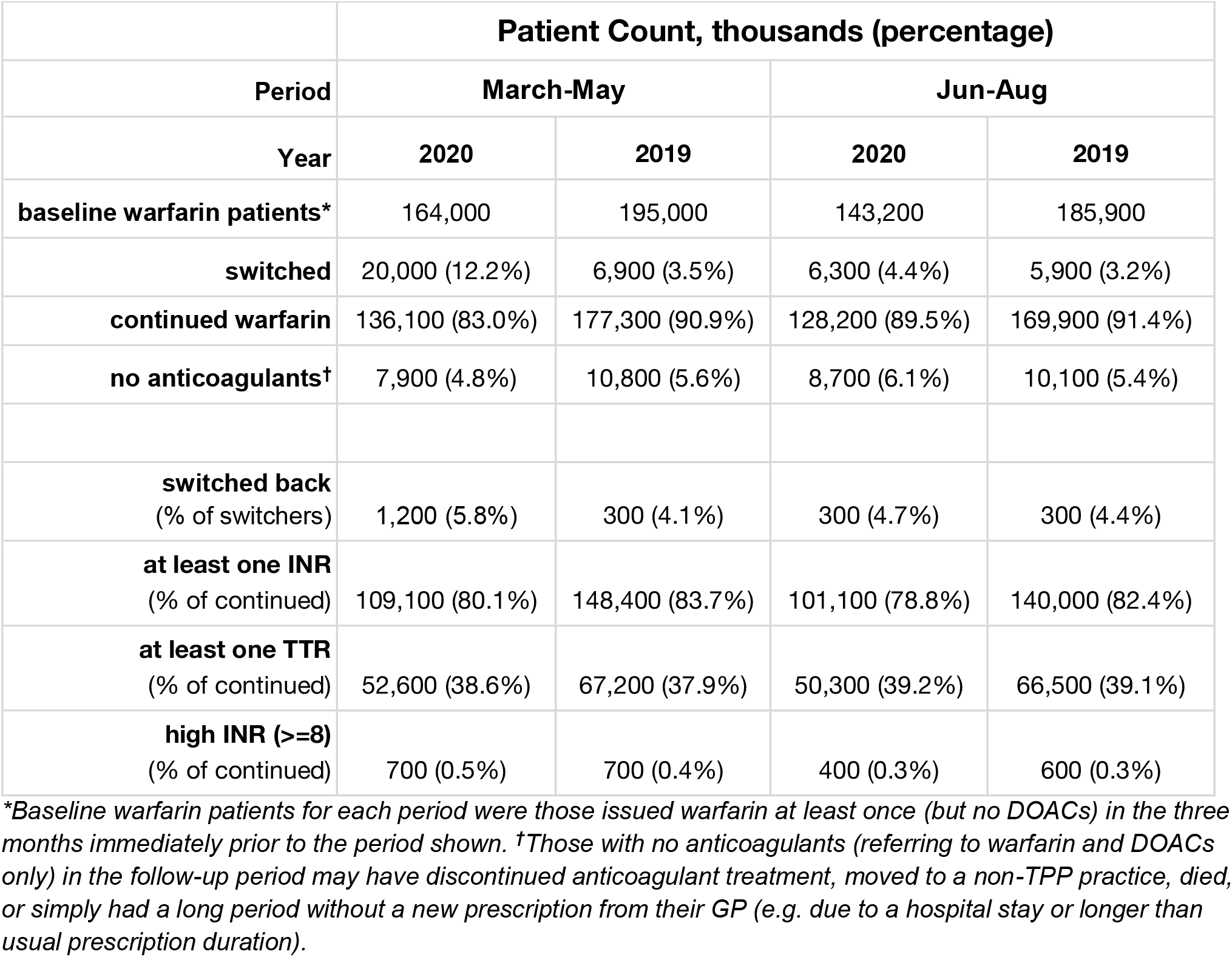
Warfarin patients switching to DOACs or remaining on warfarin. Number and percentage of warfarin patients who continued on warfarin, received a DOAC (“switched”) or had no anticoagulants, during March-May 2020 compared to 2019, and similarly for June-August. Also showing the percentage of those switching to a DOAC who later received warfarin “switched back” (within the same three month period); and the percentage of those remaining on warfarin who had at least one INR test, INR TTR, or high INR (.8) recorded. Patient counts are rounded to the nearest 100.

| Period | Patient Count, thousands (percentage) |  |  |  |
| --- | --- | --- | --- | --- |
|  | March-May |  | Jun-Aug |  |
|  | 2020 | 2019 | 2020 | 2019 |
| **baseline warfarin patients\*** | 164,000 | 195,000 | 143,200 | 185,900 |
| **switched** | 20,000 (12.2%) | 6,900 (3.5%) | 6,300 (4.4%) | 5,900 (3.2%) |
| **continued warfarin** | 136,100 (83.0%) | 177,300 (90.9%) | 128,200 (89.5%) | 169,900 (91.4%) |
| **no anticoagulants†** | 7,900 (4.8%) | 10,800 (5.6%) | 8,700 (6.1%) | 10,100 (5.4%) |
| **switched back**<br>(% of switchers) | 1,200 (5.8%) | 300 (4.1%) | 300 (4.7%) | 300 (4.4%) |
| **at least one INR**<br>(% of continued) | 109,100 (80.1%) | 148,400 (83.7%) | 101,100 (78.8%) | 140,000 (82.4%) |
| **at least one TTR**<br>(% of continued) | 52,600 (38.6%) | 67,200 (37.9%) | 50,300 (39.2%) | 66,500 (39.1%) |
| **high INR ($\geq 8$)**<br>(% of continued) | 700 (0.5%) | 700 (0.4%) | 400 (0.3%) | 600 (0.3%) |

\*Baseline warfarin patients for each period were those issued warfarin at least once (but no DOACs) in the three months immediately prior to the period shown. †Those with no anticoagulants (referring to warfarin and DOACs only) in the follow-up period may have discontinued anticoagulant treatment, moved to a non-TPP practice, died, or simply had a long period without a new prescription from their GP (e.g. due to a hospital stay or longer than usual prescription duration).

**Table 3.** Types of DOAC selected. Number and percentage of warfarin patients switched to each of the four types of DOAC between March-May 2020 compared to 2019, and similarly for June-August, for people who were prescribed warfarin during the previous three month period. Patient counts are rounded to the nearest 10. Percentages may not add to exactly 100 due to rounding.

| period | year | Apixaban |  | Edoxaban |  | Rivaroxaban |  | Dabigatran etexilate |  | Total |
| --- | --- | --- | --- | --- | --- | --- | --- | --- | --- | --- |
|  |  | patient count | % | patient count | % | patient count | % | patient count | % |  |
| March-May | 2020 | 6,700 | 33.5 | 7,620 | 38.1 | 5,550 | 27.8 | 120 | 0.6 | 19,990 |
|  | 2019 | 3,510 | 51.1 | 1,040 | 15.2 | 2,190 | 31.9 | 120 | 1.8 | 6,860 |
| June-Aug | 2020 | 2,550 | 40.3 | 2,110 | 33.4 | 1,620 | 25.6 | 40 | 0.6 | 6,320 |
|  | 2019 | 2,900 | 49.5 | 1,110 | 18.9 | 1,760 | 30 | 90 | 1.6 | 5,860 |

The initiation of repeat prescriptions for DOACs to new patients increased ∼1.5-fold during March and April 2020, and subsequently dropped slightly below normal levels (Figure 2). Much of this increase was attributable to people switching from warfarin (40.3% in March 57.5% in April), compared to the normal rate of ∼15% per month (Figure 2, Table 4).

**Table 4.** Number of people having a DOAC repeat prescription initiated per month, and the percentage of whom had previously been issued with warfarin (within the previous three months). Patient counts are rounded to the nearest 100.

| Month | Patients starting DOACs | Previously taking warfarin (%) |
| --- | --- | --- |
| Jan 20 | 11,800 | 16.2 |
| Feb 20 | 10,500 | 15.8 |
| Mar 20 | 15,700 | 40.3 |
| Apr 20 | 15,700 | 57.5 |
| May 20 | 10,200 | 38.1 |
| Jun 20 | 10,100 | 27.2 |
| Jul 20 | 9,900 | 19.8 |
| Aug 20 | 8,400 | 17.2 |
| Sep 20 | 9,500 | 15.0 |

**Table 4a.**
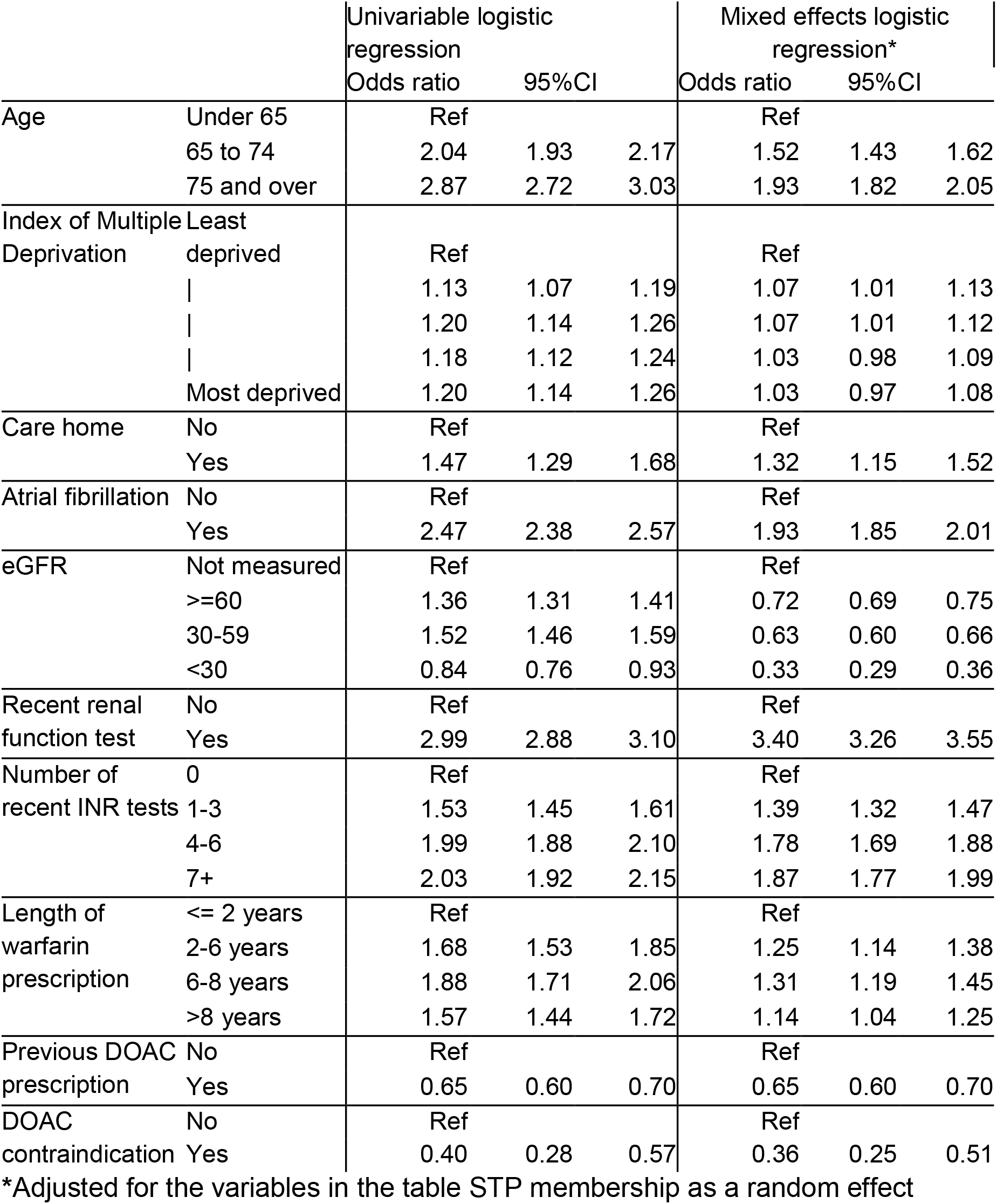
**Factors associated with switching from warfarin to a DOAC during the pandemic**

**Figure 2.**
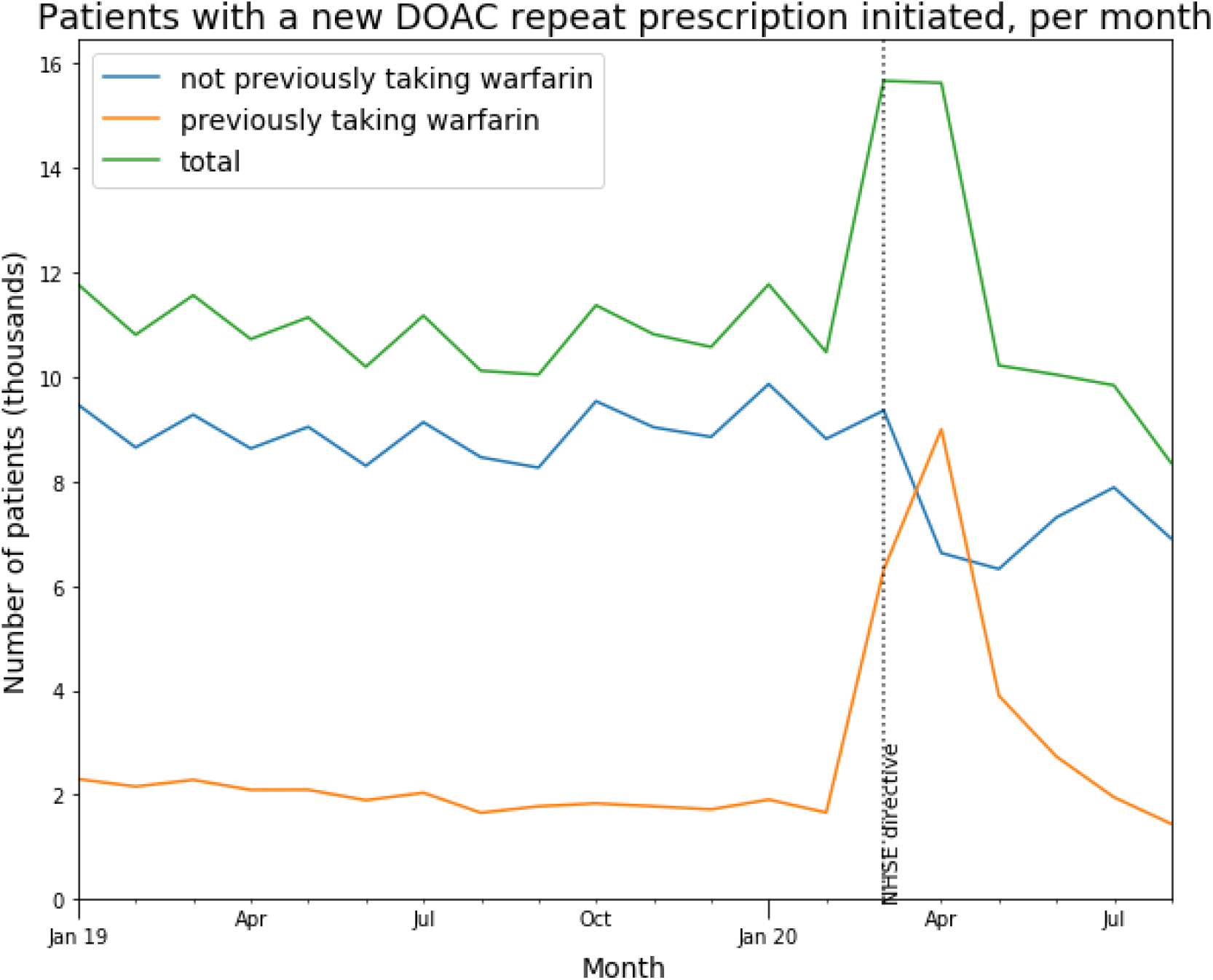
Patients newly initiated on DOAC repeats. Number of people having a new DOAC repeat prescription initiated per month (where the patient had no prior DOAC repeat ending within the previous three months), indicating whether or not patients had previously had a repeat prescription for warfarin (ending same month or within previous three months).

### Potentially unsafe co-prescribing of warfarin and a DOAC

Prior to the pandemic period, typically 50-100 people per month had both warfarin and a DOAC issued on the same day, rising sharply to 246 in April 2020 (0.06% of all people receiving a DOAC or warfarin) before declining gradually to almost baseline by August 2020 (Figure 3a). Only a small proportion of these patients had at least one of their co-prescriptions ended on the same day. Prior to the pandemic, 60-110 patients per month had both warfarin and a DOAC repeat prescription initiated on the same day (Figure 3b). This figure reached a peak of ∼170 in April, and declined rapidly to near-normal levels from May 2020. However, this rise was compensated by an increase in the number of repeat prescriptions that ended the same day (usually warfarin).

**Figure 3.**
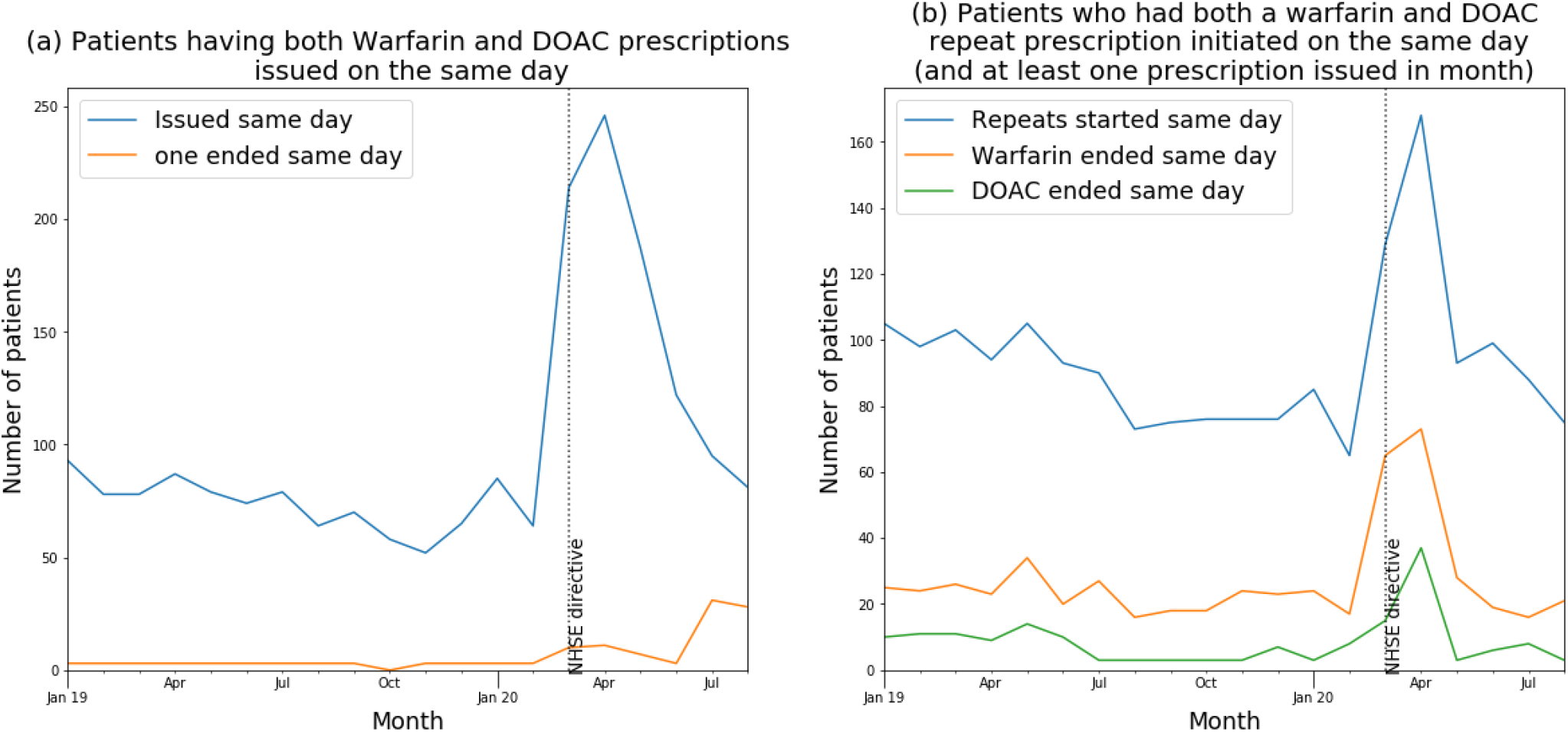
Number of patients having (a) both warfarin and a DOAC prescription issued on the same day, also indicating how many patients for whom one of those prescriptions ended the same day; (b) both warfarin and a DOAC repeat prescriptions initiated on the same day (restricted to patients who also had at least one warfarin or DOAC prescription issued in the given month), also indicating how many patients for whom one of those repeat prescriptions ended the same day.

### INR blood tests during the COVID-19 period

Of 164,000 people on warfarin prior to March 2020, 80.1% had an INR test recorded and 35% had a TTR recorded between March and May 2020 (Table 2). The number of warfarin patients tested each month was approximately constant prior to the pandemic (589 per 1000 eligible patients per month, Jan 2019-March 2020) but with a small reduction during the pandemic (Figure 4). The monthly testing rate reduced by 14%, to 506.8 (April-Aug 2020), a reduction of 82.4 per 1000 patients being tested per month (14.0% reduction). The number of TTRs recorded followed a similar pattern (Figure 5a), and where TTR was recorded the mean value for those who were still on warfarin remained relatively constant (Figure 5b). Figure 6 illustrates the rate of high INRs per 1000 warfarin patients, per 1000 INR tests and per 1000 patients tested. A small peak is observed in April, but the absolute number of high INR results (≥8) was 470, only slightly higher than January’s figure of 420 (Table S1).

**Figure 4.**
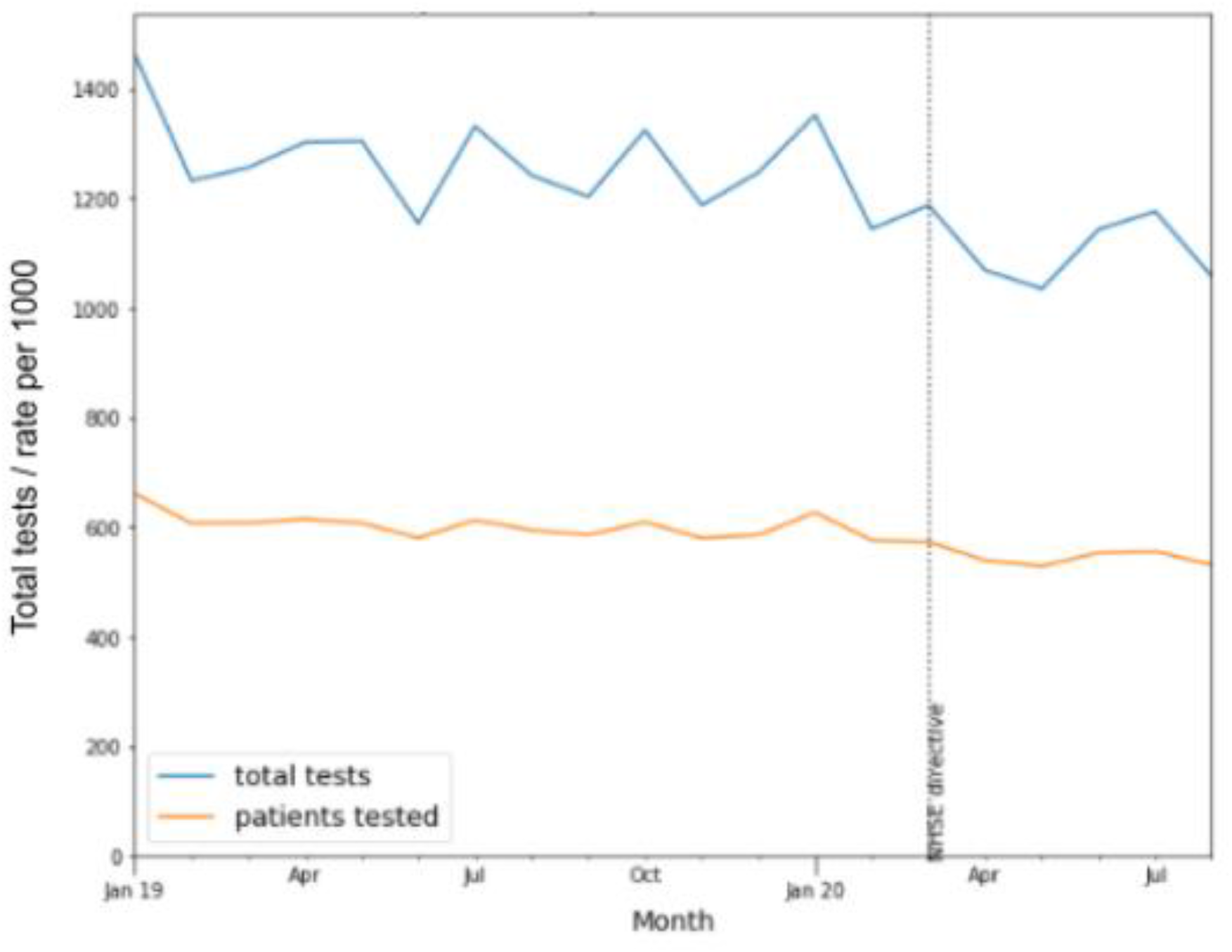
Number of patients having an INR test per month, and the rate per thousand warfarin patients.

**Figure 5:**
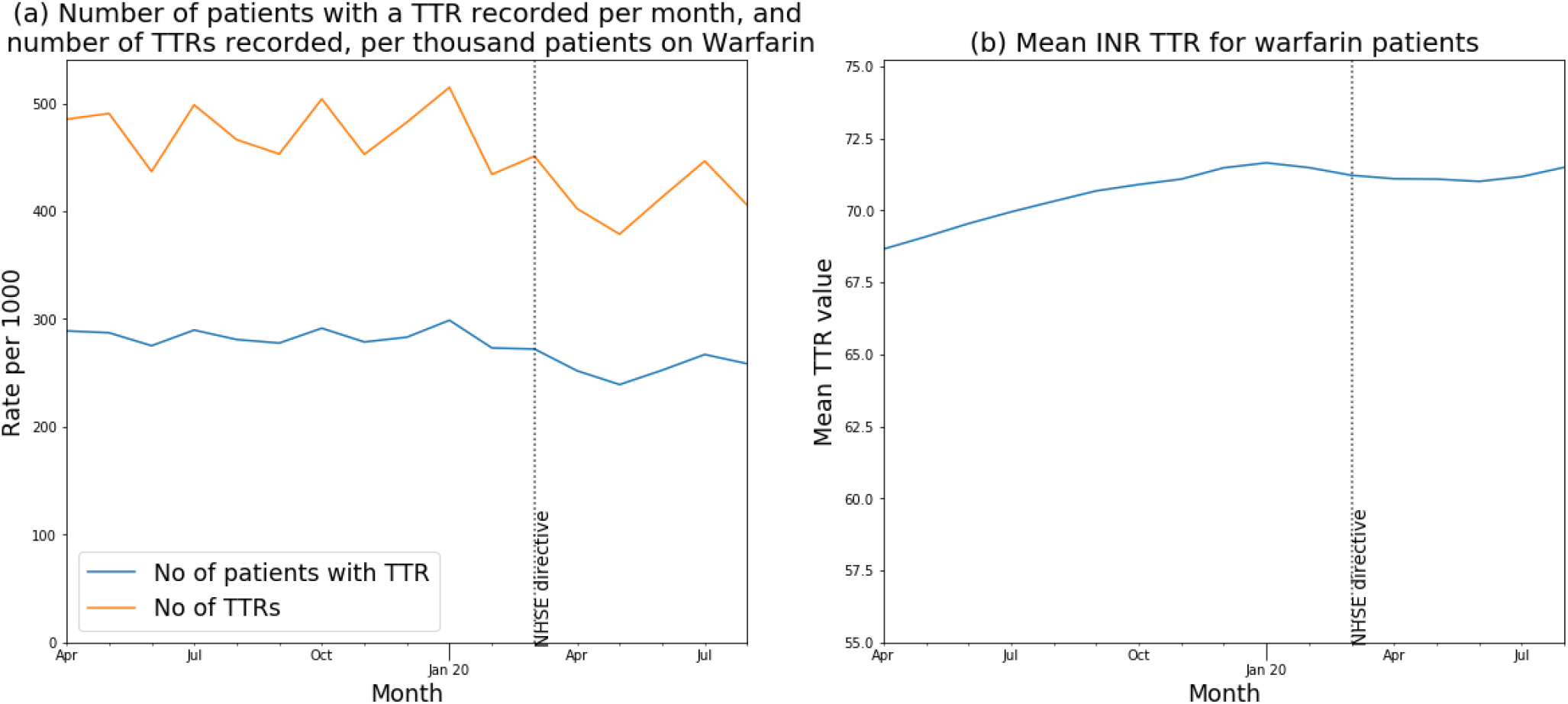
INR TTRs recorded in warfarin patients.

**Figure 6:**
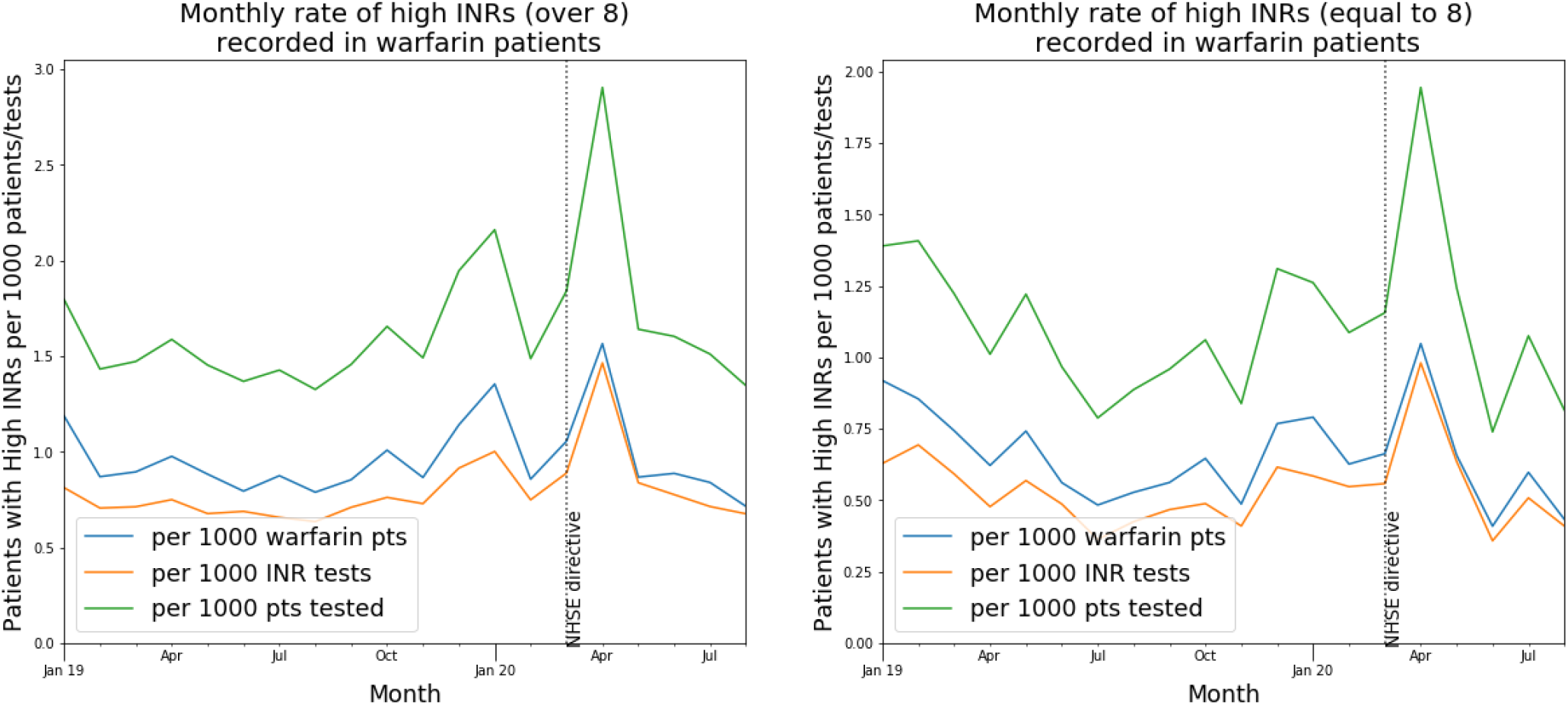
Monthly rate of elevated INRs (showing >8 or equal to 8 separately) recorded for warfarin patients relative to the total number of patients, INR tests, or patients tested.

### Factors associated with switching from warfarin to a DOAC during the pandemic

The results of our logistic regression model are presented in Table 4. People who had a recent renal function test were much more likely to switch from warfarin to DOAC (OR 3.40, 95% CI 3.26-3.55), as were people with multiple recent INR tests (over seven vs none recorded, OR 2.04, 95% CI 1.92-2.15); older people (over 75 vs under 65, OR 1.93, 95% CI 1.82-2.05); people living in a care home (OR 1.33, 95% CI 1.12 - 1.52); and people with a diagnosis of atrial fibrillation (OR 1.93, 95% CI 1.85 - 2.01). People with a recorded contraindication for a DOAC were less likely to be switched (OR 0.36, 95% CI 0.25-0.51), as were people with reduced kidney function. We also found that STP membership, when modelled as a random effect, was a significant driver of variation (p<0.0001).

## Discussion

### Summary

We observed a very substantial increase in people switching from warfarin to a DOAC after NHS England advice to do so during the COVID-19 pandemic. We were able to identify in the data that a small but substantial number of people (n=246, 0.06%) were simultaneously issued with warfarin and a DOAC in April 2020, and were potentially at risk of serious adverse effects. Overall the rate of INR testing for those on warfarin dropped by 14%, although we observed no substantial change in the proportion tests that reported a clotting time outside the desired range. Factors associated with switching from warfarin to DOACs included older age, higher number of recent INR tests, diagnosis of atrial fibrillation, recent record of a renal function test, and care home residency.

### Strengths and weaknesses

The key strength of this study is the scale, timeliness and completeness of the underlying data. The OpenSAFELY platform runs analyses across the full, raw, single-event-level medical records of all patients at 40% of all GP practices in England, including all tests, treatments, diagnostic events, and other salient clinical and demographic information. We also recognise some limitations. We assessed the number of prescriptions issued, or initiated on repeat prescriptions. We cannot currently access information on which medications were dispensed. An individual appearing to receive warfarin and a DOAC may therefore not receive, or take, both medications: for example a doctor might notice the co-prescribing some time after the consultation, and cancel the prescription before it leaves the practice; or a pharmacist may decline to dispense both medications at once to the same patient for safety reasons. Finally, a patient may be informed not to take the medicine by their healthcare professional even if they received a dispensed medicine. Nonetheless these subsequent remedial interventions do not diminish the finding that co-prescribing of warfarin and DOACs occurs, and that incidence increased substantially during COVID-19 to over two hundred people in one month. Another limitation relates to missing INR data: people in England often have their warfarin managed by an “anticoagulation clinic” in a community service, hospital outpatient department or indeed a neighbouring general practice with specialist expertise; these clinics typically use bespoke software to record all relevant results related to INR, which may not be transferred in structured data into GP records; but would commonly be visible through some other means to a clinicians delivering direct care for a patient; this could explain the apparent absence of INR tests for some people but not negatively impact clinical care.

### Findings in Context

A recent systematic review of healthcare utilisation during the pandemic found that utilisation reduced by approximately one third during the pandemic [10] but we are not aware of any published studies assessing switching of anticoagulant therapy during the pandemic or implementation of NHS England anticoagulation guidance. We found that edoxaban and apixaban were favoured DOACs when switching from warfarin, not wholly in line with NHS England advice from May 2020,[4] which recommends only apixaban and rivaroxaban. In October, NICE published draft guidance on Atrial Fibrillation recommending apixaban and dabigatran; however, we found dabigatran was prescribed in a very small proportion of switches [11]. In England, general practitioners have autonomy in the selection of treatments they prescribe, but are obliged to consider a wide number of factors which may influence choice of DOACs [12]. Our previous research has shown NHS England guidance advising GPs to stop prescribing certain medicines did not cause an immediate noticeable change in prescribing patterns [13] and CCG membership has a strong association with GP prescribing choices [14–17]. Our study shows evidence of switching after the guidance and although we were not able to directly assess CCG membership, our study did find that STPs, an administrative region made up of one or more CCGs, were a significant driver of variation. The MHRA safety alert on anticoagulation identified a “small number of patients” co-prescribed warfarin and a DOAC, but provided no further information on the scale of the problem. One Swiss hospital found 0.8% of its patients on anticoagulant therapy (121/15812) were co-prescribed two anticoagulants in a single year, 2017/2018 [18]. However, 88.7% of these cases involved co-prescription of DOACs and injectable low-weight molecular heparins, not included in our study. As regards INR blood tests, the MHRA reports appears to have been triggered by a root cause analysis from a single centre in London,[19] reporting that between 1st March and 17th April 2020, 0.9% (30/3214) of INRs were high (>8.0) compared to 0.1% (6/4079) the previous year. Analysing the full records of 40% of patients in England we found only a small peak in high INRs, and with no obvious change in the mean TTR recorded for those still on warfarin.

### Policy Implications for research and practice

The COVID-19 pandemic has brought new challenges for the NHS to deliver safe and effective routine care. NHS England issued anticoagulant guidance at the peak of the pandemic and a substantial number of people were switched in line with this guidance. This study using OpenSAFELY demonstrates it is possible to use routinely collected raw EHR data on a specific clinical area to support evaluation of national guidance and safety alerts during the COVID-19 pandemic. There is a need for high quality applied practical research to support healthcare organisations’ responses to the pandemic; OpenSAFELY can be used by NHS England, the MHRA and others such as NICE to rapidly assess in near-real time the impact of policy and clinical guidance as well as informing future versions of guidance. Specific areas of focus for anticoagulants during COVID-19 could include: identifying people eligible for switching who haven’t switched yet; assessing the scale of clinical work needed; evaluating completeness of uptake; prioritising risk groups; understanding causes of co-prescription of warfarin and DOACs; evaluating variation in organisations response to new guidance [16]; and better understanding the factors identified by MHRA as being associated with elevated INRs such as co-prescribing of antibiotics [2].

OpenSAFELY is now using full patient records to measure and mitigate the indirect health impacts of Covid-19. Our aim is to give early warning on clinical work displaced, such as cancer referrals, cardiovascular management, and vaccinations; and to monitor clinical impact on patients. We have titled this work the *OpenSAFELY NHS Service Restoration Observatory*. We can help identify NHS organisations in need of additional support as the pandemic evolves; and rapidly identify success stories from new best practices that others can learn from. Working in collaboration with NHS England and other stakeholders, short data reports will be available soon on www.OpenSAFELY.org assessing individual clinical areas and making recommendations such as research questions for examination by the research community, findings that require further analysis and actionable insights to inform care at a national, local and practice level.

### Summary

We observed increased switching of anticoagulants from warfarin to DOACs at the outset of the COVID-19 pandemic in England. We did not find a widespread rise in elevated INR test results which may be reassuring after a recent MHRA safety alert although we did observe a small but substantial number of people who were co-prescribed warfarin and DOACs.

## Data Availability

All code for the OpenSAFELY platform, and for data management and analyses for this study, are available for inspection and reuse under open licenses on GitHub (https://github.com/opensafely/anticoagulant-switching-research). All codelists are available for inspection and re-use from https://codelists.opensafely.org/.
Patient

## Acknowledgements

We are very grateful for all the support received from the TPP Technical Operations team throughout this work, and for generous assistance from the information governance and database teams at NHS England / NHSX.

## Information governance and ethical approval

NHS England is the data controller; *TPP are the data processors* and the key researchers on OpenSAFELY are acting on behalf of NHS England. This implementation of OpenSAFELY is hosted within the TPP environment which is accredited to the ISO 27001 information security standard and is NHS IG Toolkit compliant;[20,21] patient data has been pseudonymised for analysis and linkage using industry standard cryptographic hashing techniques; all pseudonymised datasets transmitted for linkage onto OpenSAFELY are encrypted; access to the platform is via a virtual private network (VPN) connection, restricted to a small group of researchers; the researchers hold contracts with NHS England and only access the platform to initiate database queries and statistical models; all database activity is logged; only aggregate statistical outputs leave the platform environment following best practice for anonymisation of results such as statistical disclosure control for low cell counts.[22] The OpenSAFELY research platform adheres to the data protection principles of the UK Data Protection Act 2018 and the EU General Data Protection Regulation (GDPR) 2016. In March 2020, the Secretary of State for Health and Social Care used powers under the UK Health Service (Control of Patient Information) Regulations 2002 (COPI) to require organisations to process confidential patient information for the purposes of protecting public health, providing healthcare services to the public and monitoring and managing the COVID-19 outbreak and incidents of exposure; this sets aside the requirement for patient consent.[23] Taken together, these provide the legal bases to link patient datasets on the OpenSAFELY platform. GP practices, from which the primary care data are obtained, are required to share relevant health information to support the public health response to the pandemic, and have been informed of the OpenSAFELY analytics platform.

This study was approved by the Health Research Authority (REC reference 20/LO/0651) and by the LSHTM Ethics Board (reference 21863).

## Guarantor

BG is guarantor.

## Supplementary

**Table S1.**
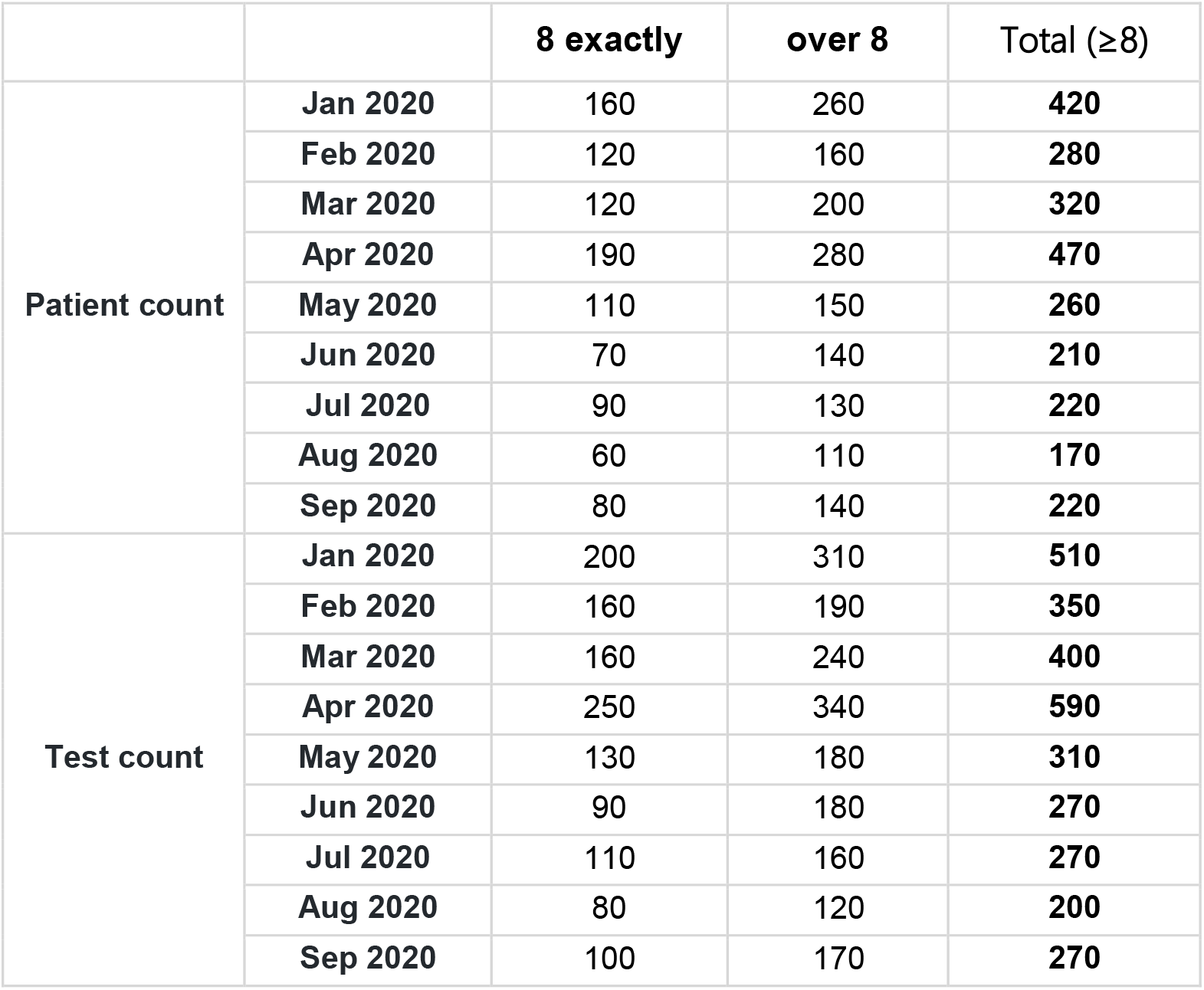
High INRs. Patient count and test count for high INR results per month of 2020, rounded to the nearest 10.

**Table S2.** Co-prescribing. Patients issued warfarin and DOAC scripts on the same day. Values below 5 are shown as “1-5”.

| **Month** | **Warfarin and DOAC issued same day** | **One ended same day** |
| --- | --- | --- |
| Jan 2019 | 93 | 1-5 |
| Feb 2019 | 78 | 1-5 |
| Mar 2019 | 78 | 1-5 |
| Apr 2019 | 87 | 1-5 |
| May 2019 | 79 | 1-5 |
| Jun 2019 | 74 | 1-5 |
| Jul 2019 | 79 | 1-5 |
| Aug 2019 | 64 | 1-5 |
| Sep 2019 | 70 | 1-5 |
| Oct 2019 | 58 | 0 |
| Nov 2019 | 52 | 1-5 |
| Dec 2019 | 65 | 1-5 |
| Jan 2020 | 85 | 1-5 |
| Feb 2020 | 64 | 1-5 |
| **Mar 2020** | **214** | 10 |
| **Apr 2020** | **246** | 11 |
| May 2020 | 187 | 7 |
| Jun 2020 | 122 | 1-5 |
| Jul 2020 | 95 | 31 |
| Aug 2020 | 81 | 28 |

**Table S3:**
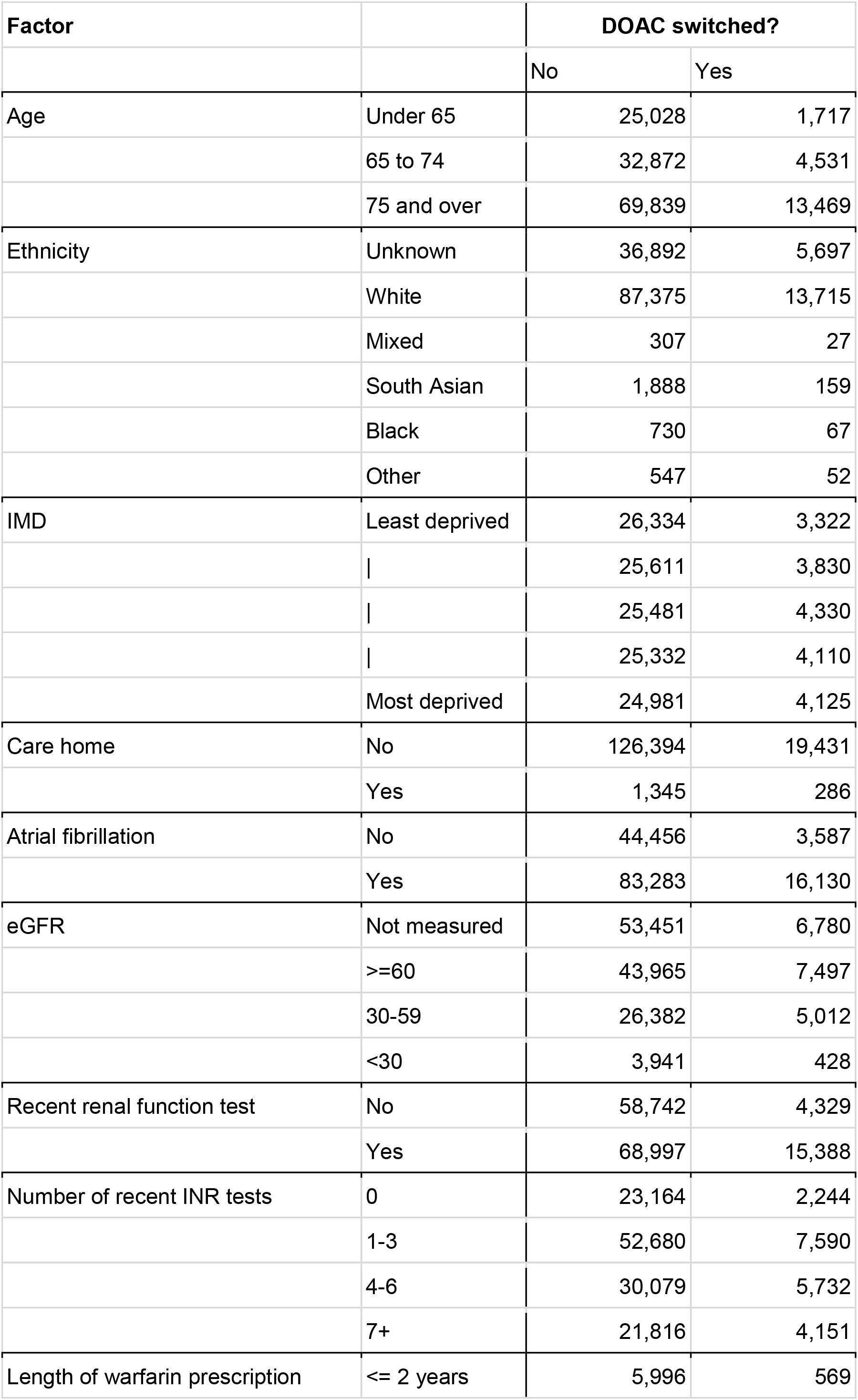
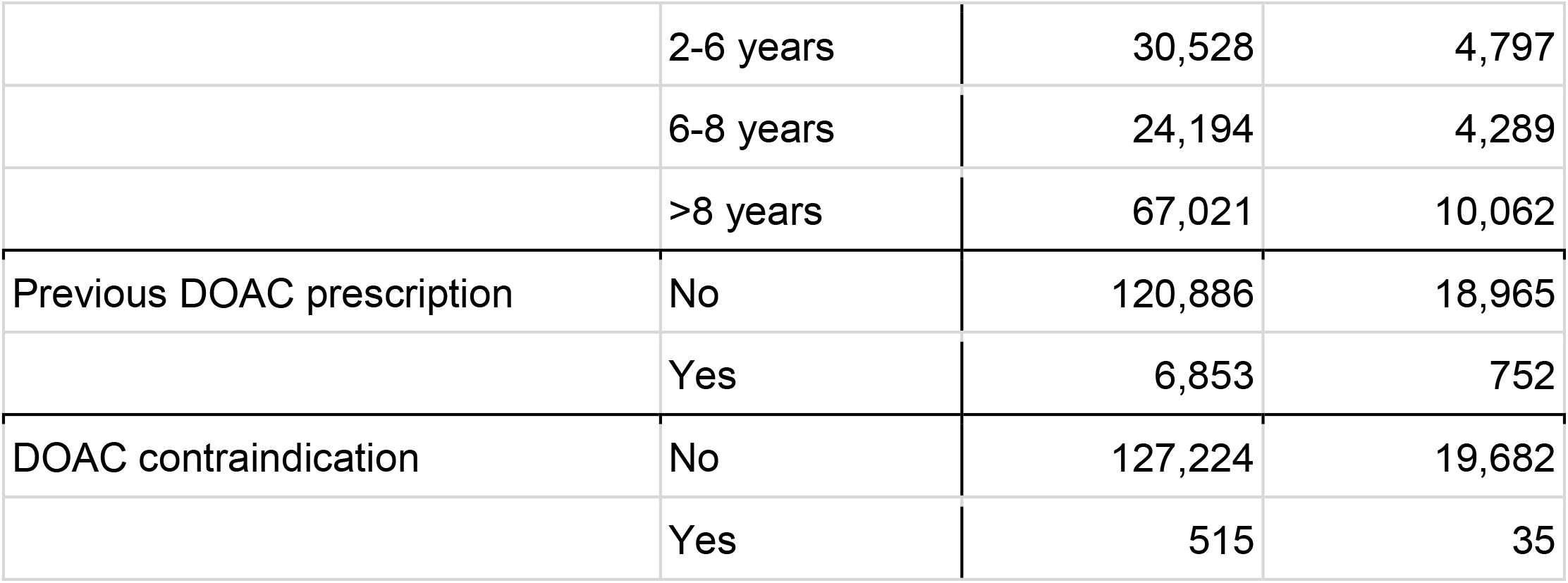
**Number of patients included in regression for factors associated with switching from warfarin to a DOAC during the pandemic, stratified by the outcome of whether the patient was switched to a DOAC**.

| Factor |  | DOAC switched? |  |
| --- | --- | --- | --- |
|  |  | No | Yes |
| Age | Under 65 | 25,028 | 1,717 |
|  | 65 to 74 | 32,872 | 4,531 |
|  | 75 and over | 69,839 | 13,469 |
| Ethnicity | Unknown | 36,892 | 5,697 |
|  | White | 87,375 | 13,715 |
|  | Mixed | 307 | 27 |
|  | South Asian | 1,888 | 159 |
|  | Black | 730 | 67 |
|  | Other | 547 | 52 |
| IMD | Least deprived | 26,334 | 3,322 |
|  |  | 25,611 | 3,830 |
|  |  | 25,481 | 4,330 |
|  |  | 25,332 | 4,110 |
|  | Most deprived | 24,981 | 4,125 |
| Care home | No | 126,394 | 19,431 |
|  | Yes | 1,345 | 286 |
| Atrial fibrillation | No | 44,456 | 3,587 |
|  | Yes | 83,283 | 16,130 |
| eGFR | Not measured | 53,451 | 6,780 |
|  | >=60 | 43,965 | 7,497 |
|  | 30-59 | 26,382 | 5,012 |
|  | <30 | 3,941 | 428 |
| Recent renal function test | No | 58,742 | 4,329 |
|  | Yes | 68,997 | 15,388 |
| Number of recent INR tests | 0 | 23,164 | 2,244 |
|  | 1-3 | 52,680 | 7,590 |
|  | 4-6 | 30,079 | 5,732 |
|  | 7+ | 21,816 | 4,151 |
| Length of warfarin prescription | <= 2 years | 5,996 | 569 |
|  | 2-6 years | 30,528 | 4,797 |
|  | 6-8 years | 24,194 | 4,289 |
|  | >8 years | 67,021 | 10,062 |
| Previous DOAC prescription | No | 120,886 | 18,965 |
|  | Yes | 6,853 | 752 |
| DOAC contraindication | No | 127,224 | 19,682 |
|  | Yes | 515 | 35 |

